# Body mass index trajectories from birth to adolescence and their association with asthma: a longitudinal study of the Leicester Respiratory Cohorts

**DOI:** 10.64898/2026.08.31.26361659

**Authors:** Lorenz M. Leuenberger, Fabiën N. Belle, Mari Sasaki, Myrofora Goutaki, Ben D. Spycher, David K.H. Lo, Erol A. Gaillard, Claudia E. Kuehni

**Affiliations:** Institute of Social and Preventive Medicine, University of Bern, Switzerland; Graduate School for Health Sciences, University of Bern, Switzerland; Department of Paediatrics, Inselspital, Bern University Hospital, University of Bern, Bern, Switzerland; Department of Respiratory Sciences, Leicester NIHR Biomedical Research Centre (Respiratory theme), University of Leicester, Leicester, UK; Department of Paediatric Respiratory Medicine, Leicester Children’s Hospital, University Hospitals Leicester, Leicester, UK

## Abstract

**INTRODUCTION:** Asthma has been associated with obesity in both adults and children. We investigated the association between longitudinal BMI trajectories and asthma at ages 8–9, 12–13, and 16–17 years in a UK cohort of White and South Asian children and adolescents.

**METHODS:** We analysed data from the Leicester Respiratory Cohorts, population-based cohort studies that recruited 0–4-year-old children in 1990 (N = 1650) and 1998 (N = 8700) and followed them up until 2010. Outcome data on asthma came from postal questionnaires throughout infancy, childhood, and adolescence. BMI data came from birth records, well-child visits, questionnaires, and a clinical study visit. We used Group-based trajectory modelling to identify distinct BMI trajectories and used multivariable logistic regression to investigate the association with asthma.

**RESULTS:** Out of 10,350 participants in the Leicester Respiratory Cohorts, we were able to model BMI trajectories for 5571 (54%); 1801 (17%) had information on asthma at 8–9 years of age, 1269 (12%) at 12–13 years, and 575 (6%) at 16-17 years. We identified five BMI trajectories: stable normal BMI (47%), persistent low BMI (30%), early overweight resolving (8%), childhood onset obesity (4%) adolescent onset overweight (11%). The persistent low BMI trajectory was associated with lower odds for asthma at age 8–9 years (0.56 [0.37–0.83]) and 12–13 years (0.38 [0.22–0.63]). Early overweight resolving was very similar to the reference stable normal BMI. The childhood onset obesity trajectory was associated with higher odds for asthma at 16–17 years (5.58 [1.35–24.40]). The adolescent onset overweight trajectory was not associated with asthma. When analysed separately, boys and girls and children of European and South Asian ancestry showed similar associations.

**CONCLUSION:** This study strengthens the current understanding of obesity as a contributing factor in asthma development and highlights that early intervention and management of overweight and obesity in childhood, to achieve a normal BMI, may prevent secondary health impairments such as asthma.

## Dear Editor

Asthma has been associated with obesity in both adults and children. Obesity related low-grade inflammation and a mismatch in lung growth resulting in dysanapsis are pathophysiological mechanisms contributing to asthma development [1-3]. Few studies have analysed how distinct body mass index (BMI) trajectories through childhood influence asthma development. Wang et al., analysing BMI trajectories from birth to early adulthood, found evidence of bronchial obstruction in young adults with persistent high or increasing BMI trajectories, but not in those with a high initial BMI that resolved later in childhood, suggesting a potential window for intervention [4]. However, the authors highlighted that results required external validation and, as they were derived exclusively from children of European ancestry in Sweden, might not be generalisable to other populations. We therefore investigated the association between longitudinal BMI trajectories and asthma at ages 8–9, 12–13, and 16–17 years in a UK cohort study that includes a large proportion (26%) of children of South Asian ancestry. We have previously reported detailed methods and results relating to BMI trajectories and their risk factors, but not their association with asthma [5].

We analysed data from the Leicester Respiratory Cohorts, population-based cohort studies investigating respiratory health [6-9]: In 1990 (N = 1650) and 1998 (N = 8700), 0–4-year-old children were randomly selected from the birth registry of Leicestershire, UK, and followed-up until 2010. Outcome data on asthma was collected from up to six postal questionnaires throughout infancy, childhood, and adolescence. Height and weight data for BMI came from three sources: birth records and well-child visits in the Leicester Health Authority Child Health Database (age 0–15 years); postal questionnaires (0–18 years); and a clinical study visit (approximately at 8 years). We used demographic, socioeconomic, perinatal, and behavioural covariates of the family from birth records and postal questionnaires.

To investigate the association of BMI trajectories and asthma, we used logistic regression. We estimated two models for asthma at each timepoint: i) univariable and ii) multivariable adjusted for demographic, socioeconomic, perinatal, and behavioural factors. We also analysed separately boys and girls, and children of South Asian and European ancestry. We accounted for the uncertainty of class membership from the trajectory modelling and used inverse probability weighting to reduce selection bias arising from differences between included and excluded participants [5].

As outcome, we defined asthma at age 8–9, 12–13, and 16–17 years from questionnaires. At each timepoint, asthma was defined as i) reported history of doctor diagnosed asthma AND ii) reported history of wheezing or use of asthma medications in the previous 12 months (short- or long-acting β_2_-agonists, inhaled corticosteroids, leukotriene receptor antagonists, or oral corticosteroids) [9].

As exposure, we modelled BMI trajectories from 0–17 years of age for participants with at least three BMI recordings, requiring at least one measured at 0–1 year and one at 3–10 years. We excluded implausible BMI values with UK-WHO z-scores <-5 or >5. We used group-based trajectory modelling with 3^rd^ degree splines to flexibly model the age pattern of BMI and specified models with up to seven trajectories. We selected the optimal model based on model fit, class size, posterior probabilities, entropy, and biological plausibility. More details are in the previous publication [5].

Out of 10,350 participants in the Leicester Respiratory Cohorts, we were able to model BMI trajectories for 5571 (54%); 1801 (17%) had information on asthma at 8–9 years of age, 1269 (12%) at 12–13 years, and 575 (6%) at 16-17 years. Of 2265 with information on asthma at any age, 52% were male, 74% were of White and 26% of South Asian ethnicity, 43% had parents with compulsory education only, 16% were exposed to maternal smoking during pregnancy, 63% were breastfed, and 43% were exposed to second hand smoke in the household.

The best group-based trajectory model identified five BMI trajectories (Figure 1): Half of the participants (47%) were in the stable normal BMI trajectory that closely followed the 50^th^ centile of the UK-WHO growth charts. The persistent low BMI trajectory (30%) followed the 40^th^ centile until 4 years of age and the 20^th^ centile thereafter, never reaching underweight levels (2^nd^ centile). The early overweight resolving trajectory (8%) was above the 91^st^ centile (overweight) from 0–13 years and resolved towards the 50^th^ centile at 18 years. The childhood onset obesity trajectory (4%) increased at 2–4 years and reached obesity levels (98^th^ centile) at 7 years. The adolescent onset overweight trajectory (11%) increased between 4–6 years and progressed to overweight at 11 years.

**Figure 1.**
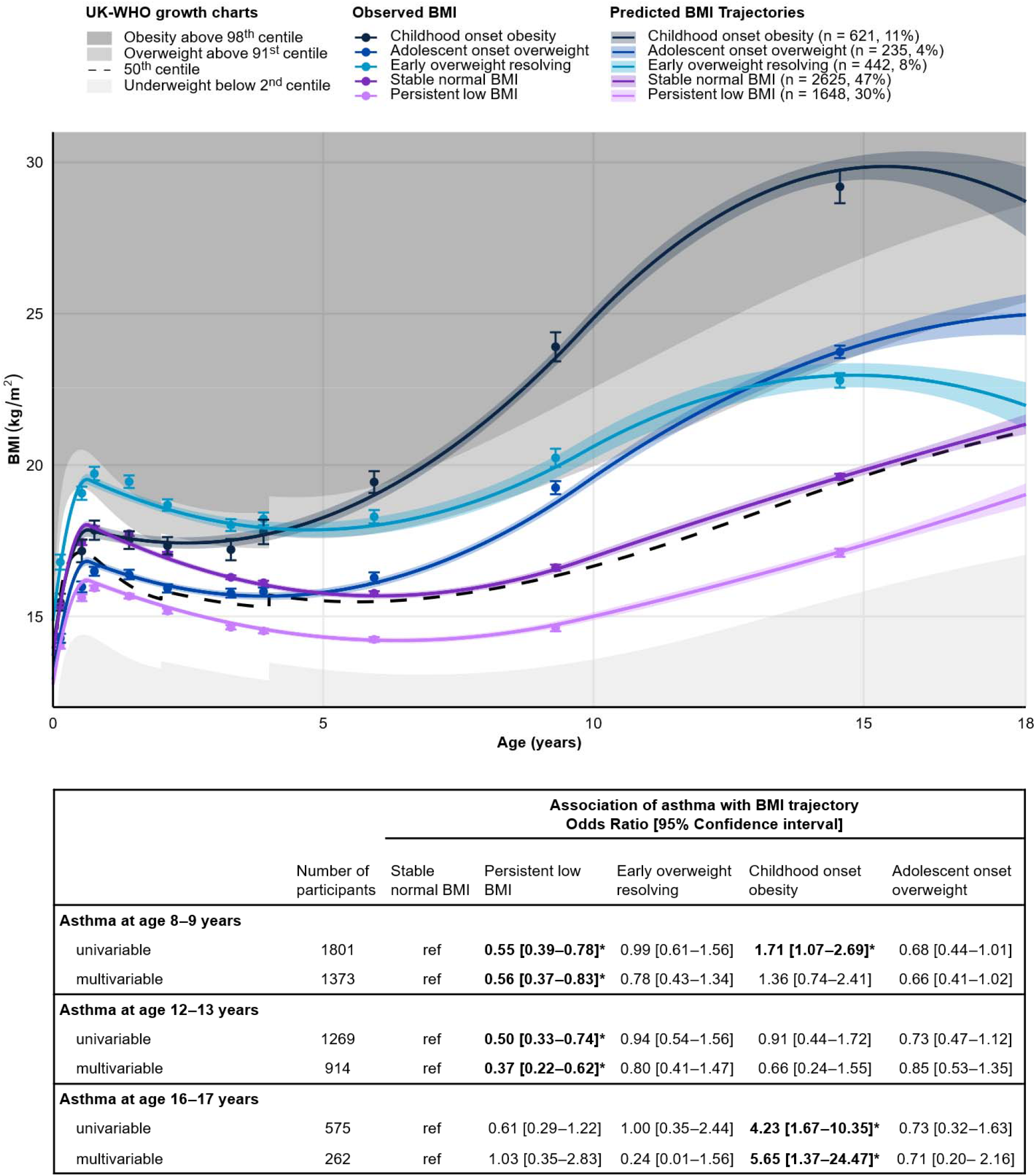
BMI trajectories of 5571 participants of the Leicester Respiratory Cohorts from 0–17 years and their association with asthma at 8–9, 12–13, and 16–17 years. BMI trajectories: Shown are mean observed and predicted BMI trajectories with their 95% confidence intervals. Associations with asthma: Multivariable models were adjusted for demographic factors (sex, ethnicity), socioeconomic factors (parental education, degree of urbanisation, Townsend index of socioeconomic deprivation), perinatal factors (birth order, maternal age at delivery, gestational age, birth weight), and behavioural factors (maternal smoking during pregnancy, breast feeding, and exposure to secondhand smoke of household members). We took the uncertainty of class membership into account by weighting each participant’s contribution to the regression model by their posterior probability of membership for each of the five BMI trajectories. We used inverse probability weighting to reduce selection bias arising from differences between participants included vs excluded in this analysis regarding the following characteristics: Sex, birth year, ethnicity, parental education, degree of urbanisation, and Townsend index of socioeconomic deprivation. * We interpreted p<0.05 as statistically significant. Current asthma: Previously diagnosed by doctor plus either wheezing in the last 12 months or asthma medication use in the last 12 months (short- or long-acting β2-agonists, inhaled corticosteroids, leukotriene receptor antagonists, or oral corticosteroids). Number of participants: We modelled BMI trajectories for 5571 participants with at least 3 BMI values from 0–18 years; 1801 of the participants with a BMI trajectory had information about current asthma at 8–9 years of age, 1269 at 12–13 years of age, 575 at 16– 17 years of age. Abbreviations: BMI: Body mass index; UK: United Kingdom; WHO: World Health Organization.

Asthma was prevalent in 15% (264/1801) of participants at 8–9 years of age, in 17% (210/1269) at 12–13 years, and in 13% (75/575) at 16–17 years. Associations of BMI trajectories and asthma were similar for univariable and multivariable models, with results from the multivariable models shown in Figure 1: The persistent low BMI trajectory was associated with lower odds for asthma at age 8–9 years (0.56 [0.37–0.83]) and 12–13 years (0.38 [0.22–0.63]). Early overweight resolving was very similar to the reference stable normal BMI. The childhood onset obesity trajectory was associated with higher odds for asthma at 16–17 years (5.58 [1.35–24.40]). The adolescent onset overweight trajectory was not associated with asthma. When analysed separately, boys and girls and children of European and South Asian ancestry showed similar associations.

Our main finding, that a high BMI trajectory is strongly associated with asthma in later life, confirms studies that have shown decreased FEV_1_/FVC in children with high BMI [4, 10]. This is also consistent with previous evidence for the influence of obesity in the first years of life on childhood asthma [11, 12], including causal effects [13]. In contrary, overweight in the first years of life that resolved during childhood and adolescence showed no increased odds for asthma in our study, which is consistent with Wang et al.’s finding of a normal FEV_1_/FVC in 24-year-olds, who had followed an accelerated resolving BMI trajectory [4]. We also show that persistent low BMI may be associated with lower odds for asthma. While a study in adults has indicated a u-shaped association between BMI and asthma, with increased asthma risk for BMI below 20 kg/m^2^ [14], studies in children have found a slightly higher FEV_1_/FVC in children with low normal BMI [4, 10].

Our study has limitations. Due to attrition, we were only able to analyse the association of BMI trajectories and asthma at 16–17 years in 575 of 10,350 participants. We reduced selection bias through inverse probability weighting. BMI may introduce misclassification of adiposity status, particularly in South Asians, in whom BMI may underestimate body fat more strongly than in Whites [15]. Therefore, we modelled BMI trajectories separately by ethnicity and sex, and found very similar trajectories [5]. We could not exclude possible reverse causation in the association of BMI and asthma; reported associations should not be interpreted as causal effects. Though our study was conducted between 1990 and 2010 we think the observed associations remain valid. A strength of our study is that we were able to adjust for a large set of exposures and thus reduce confounding.

In conclusion, we show that a childhood onset obesity trajectory may be associated with higher odds for asthma, while early overweight that resolves during childhood may have no effect. This strengthens the current understanding of obesity as a contributing factor in asthma development and highlights that early intervention and management of overweight and obesity in childhood, to achieve a normal BMI, may prevent secondary health impairments such as asthma.

## AUTHOR CONTRIBUTIONS

CEK and Michael Silverman designed and conducted the Leicester Respiratory cohort studies. CEK and LL developed the idea for this analysis. LML prepared the data and performed the statistical analyses. CEK, FNB, BDS, and MG supervised the statistical analysis. LML wrote the first draft of the manuscript, and all authors critically reviewed and revised it. All authors approved the final version of the manuscript. We did not use generative AI during study conception and data analysis. During manuscript writing and editing, we used standard editing tools for grammar and spell-checking (e.g. MS Word) and used translation software (DeepL translator, free tier) and AI tools (Gemini flash 3.6) for wordsmithing and refining single sentences, but not whole parts of the manuscript.

## ETHICS STATEMENT

The Leicestershire Health Authority Research Ethics Committee approved the study (Ref numbers 4867, 5005, 04/Q2501/61, 07/H0407/70), and written consent was obtained from children and parents for the clinical study visits.

## DATA AVAILABILITY

Data may be made available to investigators upon request by email to the corresponding author.

## ACKNOWLEDGEMENTS

We would like to thank all families that participated in the Leicester Respiratory Cohorts.

